# Bayesian investigation of SARS-CoV-2-related mortality in France

**DOI:** 10.1101/2020.06.09.20126862

**Authors:** Louis Duchemin, Philippe Veber, Bastien Boussau

**Affiliations:** Université de Lyon, Université Lyon 1, CNRS, Laboratoire de Biométrie et Biologie Evolutive UMR 5558, F-69622 Villeurbanne, France

**Keywords:** SARS-CoV-2, Bayesian model, France, Mixture model, Lockdown, Elections

## Abstract

The SARS-CoV-2 epidemic in France has focused a lot of attention as it has had one of the largest death tolls in Europe. It provides an opportunity to examine the effect of the lockdown and of other events on the dynamics of the epidemic. In particular, it has been suggested that municipal elections held just before lockdown was ordered may have helped spread the virus. In this manuscript we use Bayesian models of the number of deaths through time to study the epidemic in 13 regions of France. We found that the models accurately predict the number of deaths 2 to 3 weeks in advance, and recover estimates that are in agreement with recent models that rely on a different structure and different input data. In particular, the lockdown reduced the viral reproduction number by « 80%. However, using a mixture model, we found that the lockdown had had different effectiveness depending on the region, and that it had been slightly more effective in decreasing the reproduction number in denser regions. The mixture model predicts that 2.08 (95% CI: 1.85-2.47) million people had been infected by May 11, and that there were 2567 (95% CI: 1781-5182) new infections on May 10. We found no evidence that the reproduction numbers differ between week-ends and week days, and no evidence that the reproduction numbers increased on the election day. Finally, we evaluated counterfactual scenarios showing that ordering the lockdown 1 to 7 days sooner would have resulted in 19% to 76% fewer deaths, but that ordering it 1 to 7 days later would have resulted in 21% to 266% more deaths. Overall, the predictions of the model indicate that holding the elections on March 15 did not have a detectable impact on the total number of deaths, unless it motivated a delay in imposing the lockdown.

## Introduction

The World Health Organization (WHO) declared a pandemic of coronavirus disease 2019 (SARS-CoV-2) on March 11, 2020 following its spread to 114 countries (World Health Organization, 2020) with an estimated 118,000 cases at the time. In France, a first patient was diagnosed with the disease on January 24th 2020 (Bernard Stoecklin et al., 2020). By May 1st, the number of SARS-CoV-2 related deaths in France was 24,594 (French Government, 2020). On March 17 at noon, a lockdown was enforced that required a self-authorisation to leave home. This lockdown followed a series of less severe measures such as the prohibition of gatherings above 100 people (March 13) and school closures (March 14).

These measures surrounded already planned nation-wide municipal elections on Sunday March 15. With enforced distancing measures in polling stations, they were maintained, which led to criticism (Cedric Pietralunga, Alexandre Lemarié, Olivier Faye, 2020), as this could have favored the spread of the virus by increasing the number of contacts on a week-end day. It is therefore of interest to investigate whether these elections did have an effect on SARS-CoV-2 related deaths in France.

There has also been suggestions that different parts of France may have adhered to the lockdown requirements with different observance. Behaviours susceptible to favour the spread of the virus may have been more widespread in some regions than in others. In particular, newspapers reported that large numbers of people were not following the strict lockdown rules and instead spent time outside, typically on the banks of the Seine river, in Paris (Elsa Ponchon, 2020). If such differences between regions were true, one might expect to see an effect on region-wise numbers of SARS-CoV-2 related deaths. In particular, the Île-de-France (Paris) region would be expected to show higher mortality rates.

The lockdown was eventually lifted on May 11, when the authorities estimated that the epidemic was sufficiently under control. Given the importance of such a decision, it is important to assess the state of the epidemic on May 11 using several methodological approaches.

Various approaches have been used to monitor the epidemic. Most are compartmental models, which include Susceptible Infected Removed/Recovered (SIR) or Susceptible Exposed Infected Removed/Recovered (SEIR) models. Such models can be used in a deterministic framework, as in (Magal and Webb, 2020; Massonnaud et al., 2020; Roux et al., 2020; Sofonea et al., 2020), can be used for performing simulations by including stochasticity through resampling steps in an otherwise deterministic framework (Neher et al., 2020), or can be used in a completely stochastic framework, as in (Flaxman et al., 2020; Salje et al., 2020). Deterministic models have small computational requirements, but probabilistic approaches lend themselves to statistical inference, e.g. Bayesian inference.

In this paper we used Bayesian inference to study SARS-CoV-2 related deaths in France. We build upon work by Flaxman et al. (Flaxman et al., 2020) to investigate heterogeneity of the viral reproduction number *R_t_* due to both temporal (lockdown, week-ends, election day) and spatial variations (inter-regional heterogeneity), and to evaluate the status of the epidemic when the lockdown was lifted on May 11.

Flaxman et al. proposed a Bayesian method to estimate decreases of the reproduction number (*R_t_*) of the virus due to various interventions such as school closures and lockdowns among 11 countries. We adapted this model from its released version 2. Version 2 improves upon version 1 by accounting for the fact that *R_t_* decreases as the pandemic progresses because a larger portion of the population has been infected and can no longer be infected. We applied the model to the 13 French regions and notably computed region-wise Infection Fatality Rates (IFR) by taking into account region-specific demographic data. First, we investigated the ability of the model to predict the progression of the epidemic in France. Second, we examined the effect of the lockdown on the reproduction number of the disease. Third, we examined the ability of the model to detect two types of temporal heterogeneities: weekends, during which a smaller portion of workers go to work, and March 15th election day.

We used simulations to assess the effect size necessary for the model to detect these heterogeneities, and then applied the model to the empirical data. Fourth, we developed a mixture model to study potential heterogeneities among regions. We found that this model had a better fit than the first model. Fifth, we used both model 1 and the mixture model to assess the total number of infections as of May 11, and the new infections on that day. Finally, we investigated counterfactual scenarios in which the lockdown is imposed 1 to 7 days before or after the actual date.

## Methods

### Models

#### Basic model

Here we present the version 2 of the model by Flaxman et al. (Flaxman et al., 2020) briefly, and direct the interested reader to the original publication for more details. We have kept the original authors’ symbols for clarity. Version 2 models the evolution of the number of deaths day by day by assuming a discrete renewal process, where portions of the population are susceptible, infected, or recovered/dead. This process describes the evolution of the number of infections over time, and serves as an input to a model of the time between infection and death. In the original model, heterogeneities between countries were induced by different input parameter values. For instance, each country had its own population size. All the countries however shared the same estimated parameter values, apart from parameters setting the number of seed infections, which describe the numbers of infections happening during the first 6 days of the epidemic in a given country, and are necessary to initiate the epidemic. The model accounted for variations in the reproduction number of the virus due to non-pharmaceutical interventions. It estimated parameter values for each of the interventions, which were shared by all countries.

More specifically, deaths on a given day are the consequence of infections that took place some infect ion-to-death time in the past. The model allows for variation across individuals in this inf ection-to-death time by assigning it a probabilistic distribution n. In practice n is the convolution of two Gamma distributions whose parameters are obtained from the literature. That is, the inf ection-to-death time is modeled as the sum of two independent random times: the incubation period, and the time between onset of symptoms and death. Both time components are Gamma distributed. The observed daily numbers of deaths *D_t_,_m_* on day t for region m are drawn from a negative binomial distribution with parameters that vary day by day:

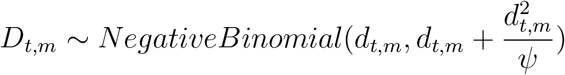

where ψ *~ Normal^+^* (0,5) is a half-Normal distribution. 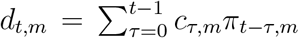 is the expected number of deaths on day *t* for region *m*. It is a discrete sum of the number of new infections *c_τ_,_m_* per day *τ* and region *m* since the first day of data, times the probability *π_t−τ_,_m_* that people infected on that day *τ* die on day *t*. The number *c_τ_,_m_* of new infections on day *τ* and region *m* is the result of a discrete renewal process. This process depends first on a distribution *g* of time between infection and the ability to infect other individuals, and second on a country-specific reproduction number *R_t_,_m_*. *g* is set to be a Gamma distribution with parameters fixed. *R_t_,_m_* models the average number of secondary infections at time *t* for country *m*. It depends on:

- the population size of the country: *R_t_,_m_* will tend to be larger in larger populations as there are more people to infect. However, as the number of infected and recovered individuals increases in a country, *R_t_,_m_* decreases because there are fewer individuals to infect. This is handled in the version 2 model deterministically based on population sizes given as input to the model.
- the age structure of the country to account for the variable susceptibility of the different age classes in a population. *R_t_,_m_* will tend to be larger in countries with older populations. This is handled in the version 2 model deterministically based on *infection fatality ratios* (IFR) given as input to the model.
- non-pharmaceutical interventions such as a lockdown. By reducing the number of contacts between individuals, these interventions will tend to reduce *R_t_,_m_*. The effect of each intervention is quantified by a single parameter that we seek to estimate from the data. It is assumed to be homogeneous over all days during which it is enforced.

#### Model extensions

Our models reproduce the general structure of the version 2 model. However we applied it to French regions, with changes in the type and number of interventions, and, in one case, allowing for different estimated parameter values for different regions.

We used four models: one model where only the lockdown is included, one model with lockdown and week-ends, one model with lockdown and election day, and one mixture model with lockdown allowing for heterogeneities among regions in the efficiency of the lockdown.

1. Model with lockdown. The model with lockdown is basically the same as in (Flaxman et al., 2020) except that a single intervention was considered. Lockdown was considered to have an homogeneous effect throughout all *m* regions and from its start to its end. It was assumed to have an effect on the reproduction number *R_t_,_m_* of the virus according to equation 1:

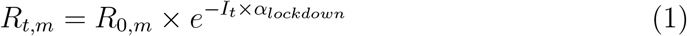

where *R*_0,_*_m_* stands for the reproduction number at day 0 in region *m* and incorporates demographic parameters, and *I_t_* stands for an indicator function for day *t* taking value 1 on lockdown days and 0 otherwise. The prior distribution of *α_lockdown_* is a Gamma distribution of mean 0.1667 and standard deviation 1.0, shifted to the left to allow for decreasing or increasing effects with about a 50/50 chance. For this intervention, large decreasing effects are expected, so the distribution was mirrored around 0 by taking its negative, leading to the prior shown in 2.

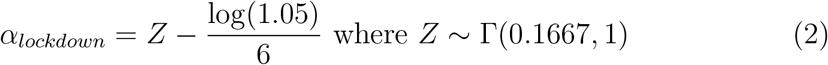
2. Model with lockdown and week-ends. The second model builds upon the first model by including the influence of week-ends. These were modelled as an additional intervention with the same prior as for the lockdown, assuming less work on week-ends compared to weekdays should induce lower reproduction numbers (Eq. 3). However, let it be clear that this model is not intended to explain the irregularities in mortality *reporting* during week-ends.

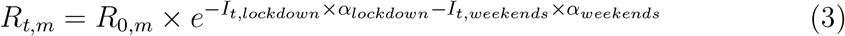
3. Model with lockdown and election day. The third model builds upon the first model and includes the influence of the election day. On this single day, another intervention is added, with a prior very similar to that used for the two other interventions, except that we expect here an *increase* of the reproduction number. Therefore, we used the same prior as for the other interventions except for the negative sign, yielding equation 4.

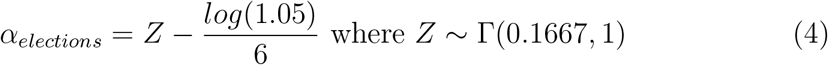

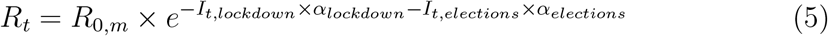
4. Model with heterogeneity among regions. The fourth model builds upon the first model but allows for heterogeneity among regions with two categories. These two categories of regions are allowed to differ in how much the lockdown changed the transmissibility of SARS-CoV-2. To this end, we implemented a mixture model on *α_lockdown_* parameters, with two categories, resulting in two parameters, 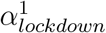 and 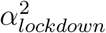. A region *m* can choose between the two possible values, and this is indicated with a Bernoulli distributed variable *C_m_* ∈{1, 2}. We called *θ* the parameter of the Bernoulli distribution, and chose a uniform prior for it. In summary:

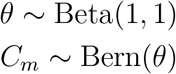 Then we defined *R_t_,_m_*, the reproduction number for region *m* as:

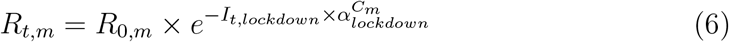 We draw both 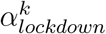 values from the same prior distributions as for the first and second models, but enforce that 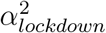 is larger than 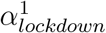 by using a dedicated variable type in Stan (Stan Development Team, 2019). Since Stan does not handle mixture models explicitly, we encoded a marginalized version of our model as proposed in Stan’s manual and developed a posterior decoding method (described in Supplementary Material 4.1) to extract results for individual regions.

### Data

#### Mortality data

Mortality data per region were downloaded on May 11 2020 from two sources: OpenCovid (OpenCOVID19 contributors, 2020), and Sante Publique France (SPF) (French Ministry of Health, 2020). OpenCovid is a citizen-based initiative, whose aim is to assemble and provide data sets to study the epidemic in France and abroad. SPF is a governmental agency that provides data related to the epidemic at national and sub-national levels. Both datasets were merged into one, prioritizing data from SPF on the days when observations from both sources were available.

Data for regions Guadeloupe, Guyane, La Réunion, Martinique, and Mayotte, which have low mortality numbers in the studied period, were not included in this analysis. The first day for which we have data in all regions is February 15. The amount of missing data from this day onward is low: 14 days at most for regions Île-de-France, Occitanie and Pays de la Loire, and 10.92 days on average (fig. 1).

**Figure 1.**
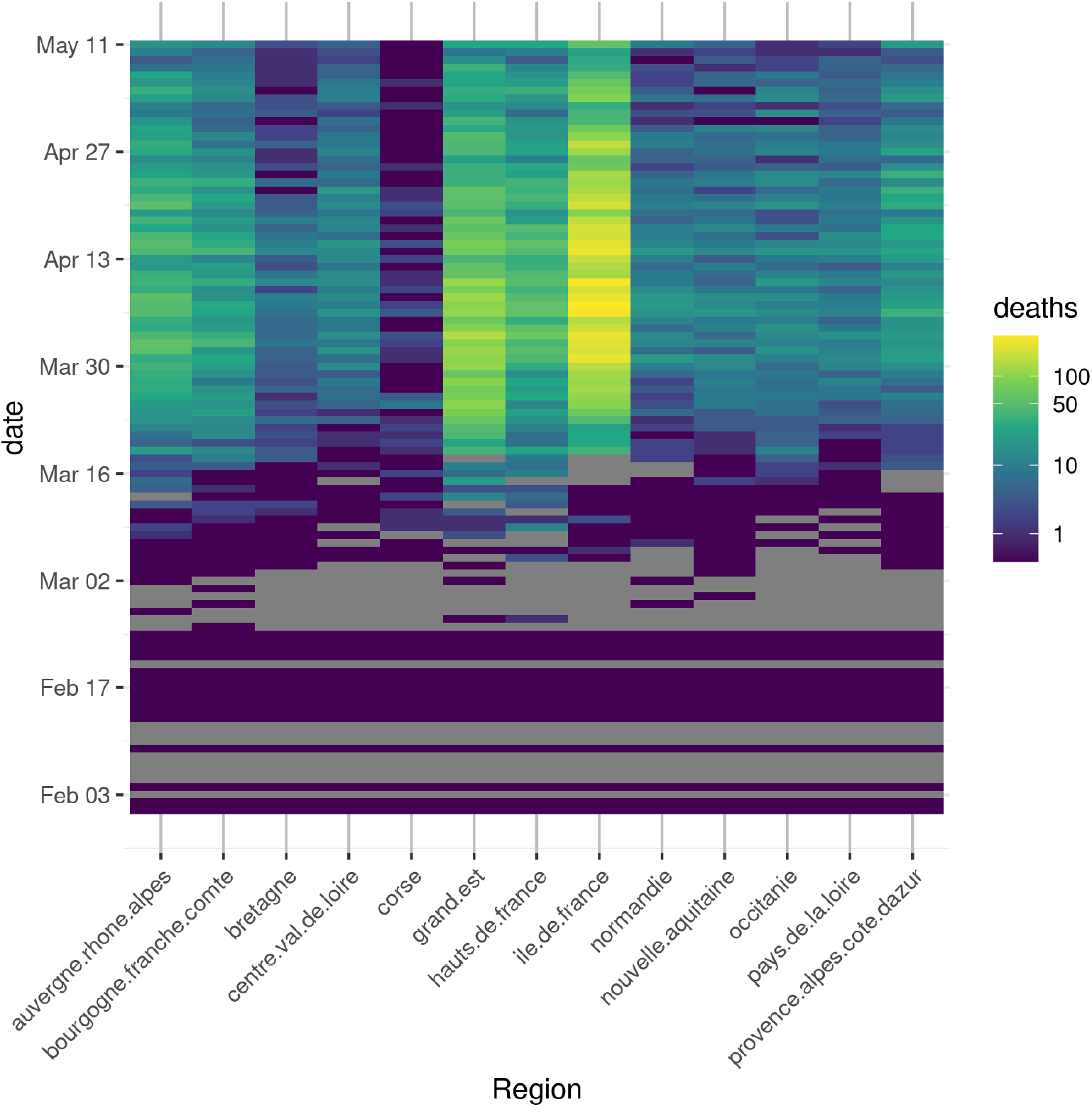
Mortality data for 13 regions in France, from the first day when all regions have data. Colors are scaled as log mortality for a given day and region. Gray tiles indicate missing data. All data from March 19th onwards originate from the SPF dataset.

#### Infection Fatality Ratios

Infection Fatality Ratios (IFRs) provide the probability of death given infection, and vary depending on the age of the infected individual. Based on data from China, IFRs were estimated for 9 age classes: [0 - 9], [10 - 19], …, [70 - 79], [80 <] by (Verity et al., 2020). Those estimates cannot be used directly for French regions as many parameters susceptible to affect IFRs differ between the two countries. However Flaxman et al. (Flaxman et al., 2020) estimated country-specific Case Fatality Rates (CFRs), providing the probability of death given a diagnosed infection. We used the country-wise CFRs for China (0.0138) and France (0.011526) to scale the Chinese age-specific IFRs. More specifically, we use proportionality to scale all Chinese age-specific IFRs by 0.011526/0.0138 to obtain French age-specific IFRs. Finally, we obtain region-wise IFRs by computing the sum of the French age-specific IFRs weighted by the population size of the corresponding age class in each region.

### Choice of interventions

In (Flaxman et al., 2020), different interventions had been used: school closure ordered, case-based measures such as self-isolation, public events banned, social distancing encouraged, lockdown decreed. In France, these different interventions happen in close temporal proximity, at the same time in all regions, between March 13 and March 17. This makes identifying their individual contributions very challenging. Therefore we chose to only use one intervention, the full lockdown, on March 17. We also considered two additional events, that were treated in the model as additional interventions: week-ends and the election day, as each could have an effect on the viral reproduction number. In particular, week-ends may decrease *R_t_* because more businesses are closed on week-ends, and the election day may increase *R_t_* by gathering many voters in polling stations.

### Simulations to estimate effect sizes

We investigated the ability of the model to detect the effect of one-day events, like the elections, or of week-ends, depending on the size of the effect.

To do so, we relied on simulations reproducing the model’s dynamics, and accounting for the effect of the events to be investigated (elections or week-ends) as described in section. Each simulation was initialized with parameters sampled from a previous fit of the model. The reference model used to sample these parameters accounted for the lockdown effect, and was fitted on mortality data up to May 11, yielding 2000 samples of parameter values. 500 sets of parameters were randomly sampled from this pool in order to run 500 simulations per conditions.

Conditions were defined as a fold-change applied to the adjusted *R_t_* during the elections or week-end days. With our prior hypotheses that week-ends would cause a decrease in *R_t_*, we ran simulations assuming fold-changes: 1 (no change), 0.9, 0.75,0.5. Similarly, to evaluate the consequences of a putative *R_t_* spike during the elections, we ran simulations with fold-changes: 1, 1.25, 1.5, 2. We then compared the simulated mortality between conditions to evaluate the possibility to retrieve such a change in *R_t_* from mortality observations.

### Implementation

The models described in paragraph were encoded using Stan’s probabilistic language (Stan Development Team, 2019), as variants of the code developed by Flaxman et al. (Flaxman et al., 2020) (version 2). Inference was performed using Stan via the R library rstan. Stan implements a variant of Markov Chain Monte Carlo (MCMC) inference algorithms, called Hamiltonian Monte Carlo (HMC). Given a model with unknown parameters and data, this algorithm generates a sequence of parameter values whose distribution converges to the posterior distribution of the parameters given the data. In our inferences, we used 4 independent chains. We discarded the initial 2000 iterations of each chain (burnin) and used the next 4000 iterations for our posterior sample. Convergence of the chains was assessed by checking the Rhat statistic which is based on comparing inter-chain to intra-chain variance, as recommended in Stan’s manual.

## Results

We first investigate whether model 1 can capture the major trends of the epidemic in the French regions. Second, we use it to evaluate the efficacy of the lockdown. Third, we study the ability of models 2 and 3 (section) to identify changes in the reproduction number due to the elections or to week-ends, both on simulated and empirical data. Fourth, we investigate potential differences among regions in the efficacy of the lockdown. Fifth, we study counterfactual scenarios where the lockdown is enforced a few days before or after March 17 to evaluate the effect on the total number of deaths.

### Evaluation of Model 1 and of the efficiency of the lockdown

#### Model fit

(Flaxman et al., 2020) investigated the fit of their model by cross validation. To do so, they pruned from their data set 3 days for which they have data and compared the inferred numbers of deaths to the empirical numbers of deaths. They repeated this procedure several times. The model was found to behave well, with a correlation of 93% between the inferred and empirical country-wise numbers of deaths. We challenged our model a bit further by predicting the number of deaths in the 13 regions of France after hiding large parts of the data. Each run was performed by removing the *k* last weeks of data, with *k* ranging from 0 to 8, and comparing the inferred and empirical numbers of deaths up to May 11 when the lockdown was lifted. Remaining data points used for estimation after removing those weeks are refered to as “prefix” in this section.

Fig. 2 shows the results when different numbers of days are given as input for region “Tle de France”. Data for other regions are presented in Supp. Mat. and show the same trends. The data shows weekly trends of low numbers of deaths on week-ends compared to high numbers just after the week-ends. This can be explained by the fact that the counts provided by French public health agencies are based on the date each event was reported, and not the date it occurred (Luc Peillon, 2020). However in practice there is always a latency between the events occuring during the treatment process (e.g hospitalization, admission in ICU, decease) and their reporting. This latency is longer during the week-ends, possibly because of reduced workforce, leading to increased numbers reported on the following Monday. The model does not explicitly handle under-reporting and instead smoothes these irregularities out.

**Figure 2.**
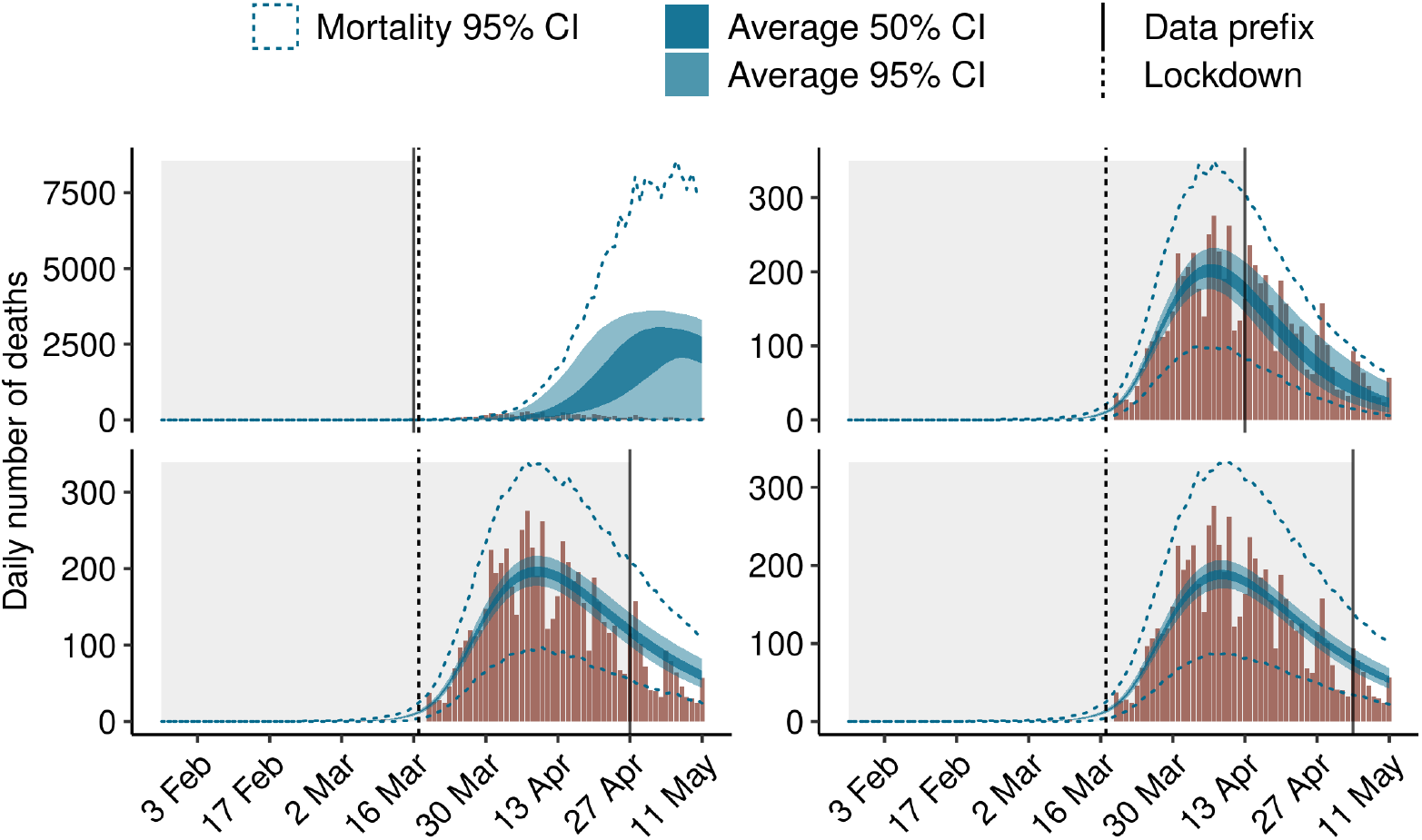
Model fits using prefixes of data for region Ile-de-France. The dashed vertical line corresponds to March 17, when the lockdown was enforced. Data right of the plain vertical line were hidden from the model. The observed numbers of deaths are represented with a brown histogram, and the predictions of the model are in blue. Dark blue ribbons correspond to the 50% credibility intervals and light blue ribbons to the 95% credibility intervals of the expected numbers of death. Blue dashed lines represent the 95% credibility interval of the predicted numbers of deaths *D_t_,_m_* (see section).

The model both predicts the expected numbers of deaths per day and the actual numbers of deaths, which are simulated thanks to a negative binomial distribution around the expected numbers of deaths. The model performs poorly when the last 8 weeks of data are held out (upper left panel), and vastly overestimates the numbers of deaths. This is likely due to the fact that with such an early censoring of the data, no information about the lockdown is given to the model. The three other panels show that when 4 or more additional weeks of data are provided, the model does a good job at predicting the dynamics of the epidemic. These 4 additional weeks provide the data necessary for the model to estimate the effect of the lockdown on the reproduction number.

For instance, the model estimates that in total there had been 6231 deaths [CI: 5456-7160] in region “Tle de France” when all the data up to May 11 is used, 6502 deaths [CI: 5698-7403] when the data stops one week before May 11 (bottom right panel), 6829 deaths [CI: 5908-7882] when the data stops two weeks before May 11 (bottom left panel), and 5894 deaths [CI: 4854-7443] when the data stops four weeks before May 11 (top right panel). The actual total number of deaths on May 11 in this region is 6643, which is in the credibility interval for all estimates.

To focus on the predictive ability of the model, i.e. its ability to estimate the number of deaths for unobserved weeks, we computed the total squared error only on the last unobserved week of data, and varied the prefix size. With a prefix that stops right before this last week, the total squared error is 12350 (95% CI: 7051-25307). If the prefix stops 2 weeks before the last week, it is 14956 (95% CI: 8036;35293), and 18001 (95% CI: 11420;27495) if the prefix stops four weeks before the last week. The error made by the model when predicting 4 weeks in advance is thus 45% worse than when predicting one week in advance. We conclude from the above that the model can be used to predict the number of deaths several weeks in advance while keeping a useful level of accuracy.

Figure 3 presents fitted mortality for three regions, using data up to May 11. Equivalent figures for all regions in this analysis are provided as supplementary material.

**Figure 3.**
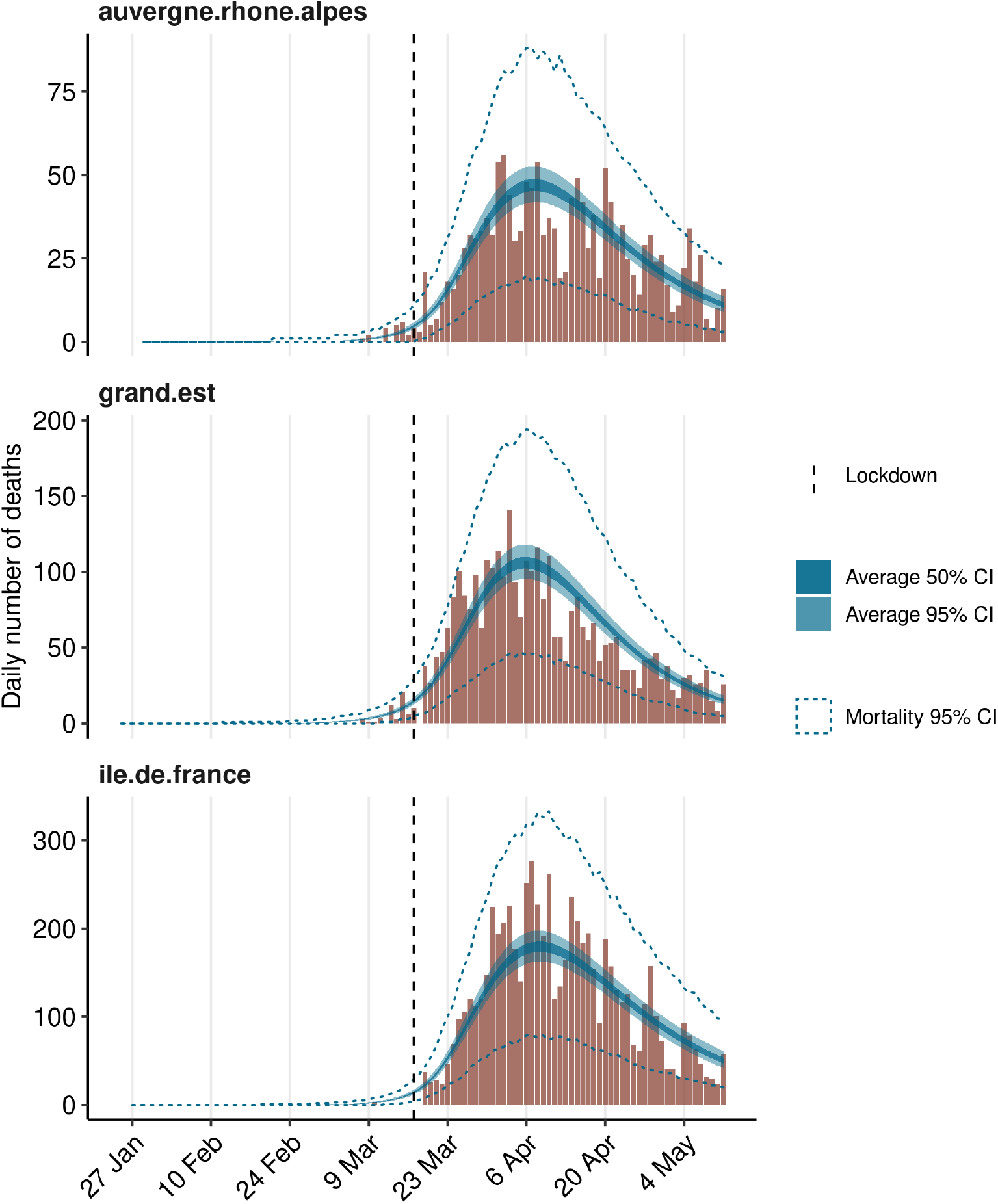
Model fit on the complete dataset for three different regions.

If we focus on the total number of deaths in France using data up to May 11, we observe that the model is able to reproduce the trends in the observed numbers very accurately, making errors ranging from 0.86% (9750 estimated deaths for 9834 observed in data) to 6.70% (7300 estimated for 7824 observed) per day over the month of April (Fig. 4). This shows that the inability of the model to capture weekly irregularities in the reporting of deaths has not had a noticeable effect on the estimation of the total numbers of deaths through time.

**Figure 4.**
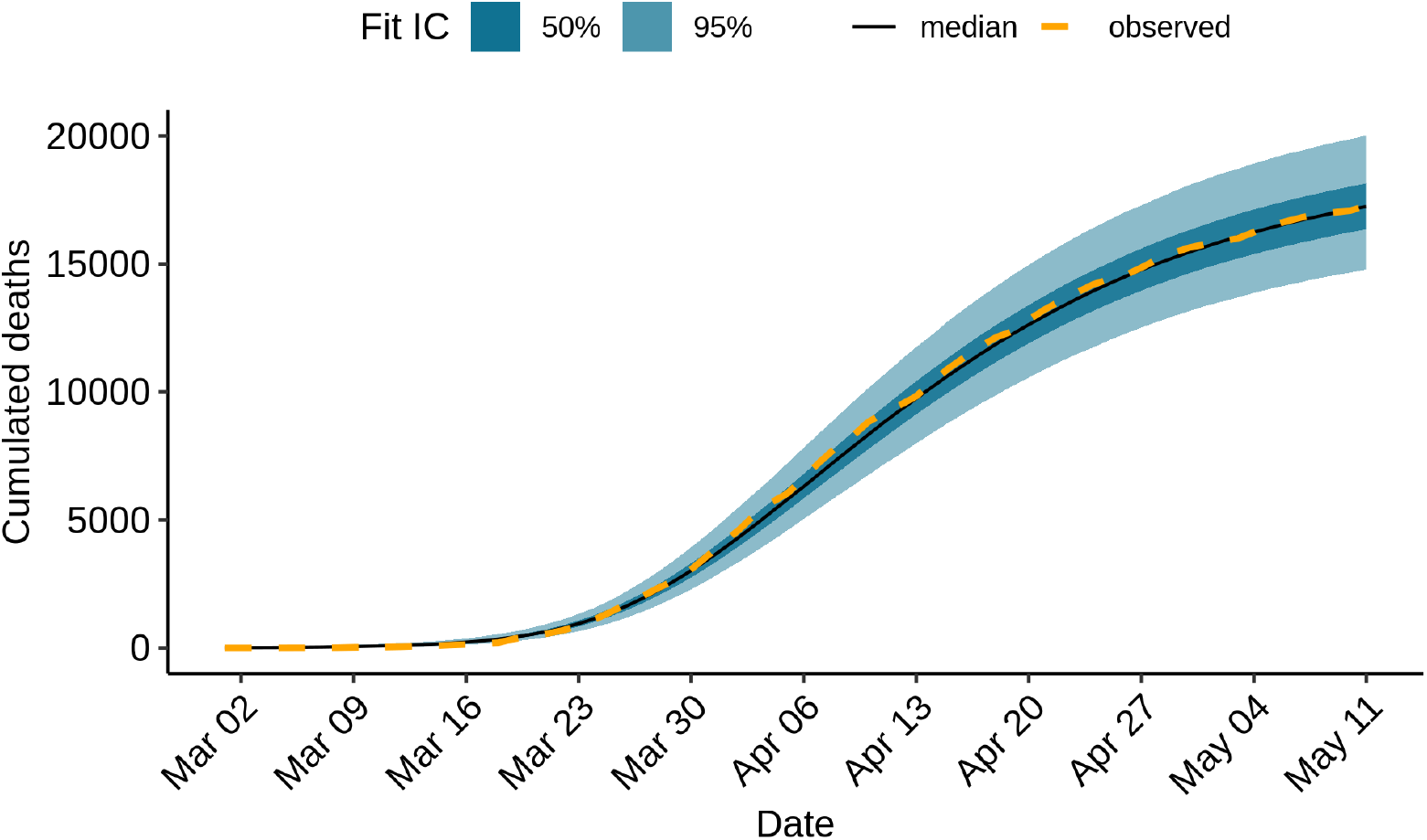
Cumulated mortality over time, fitting data up to May 11.

Overall, the model appears to capture well the dynamics of the epidemic in French regions. In the following, we use the model to investigate whether particular events in the pandemics in France have left a footprint in the number of deaths.

#### Reduction of viral transmissibility due to the lockdown

Model 1 allows estimating the effect of the lockdown on the reproduction number of the virus. This is done through a parameter *α_lockdown_* whose prior distribution is a shifted Gamma (see section). The posterior distribution clearly differs from the prior distribution meaning that there is information in the data to estimate the *α_lockdown_* parameter value (Supplementary Figure 11).

As shown Fig. 5, the reproduction number in Ile-de-France decreases markedly with the lockdown, shifting from about 3.58 (95% CI: 3.34 - 3.86) before the lockdown to 0.69 (95% CI: 0.65 - 0.73) after the lockdown, *i.e*. a reduction of 80.78%.

At the national level, the average *R_t_* among regions weighted by their population size is 3.34 (95% CI: 3.19 - 3.51) before lockdown and decreasing to 0.65 (95% CI: 0.62, 0.67) after.

**Figure 5.**
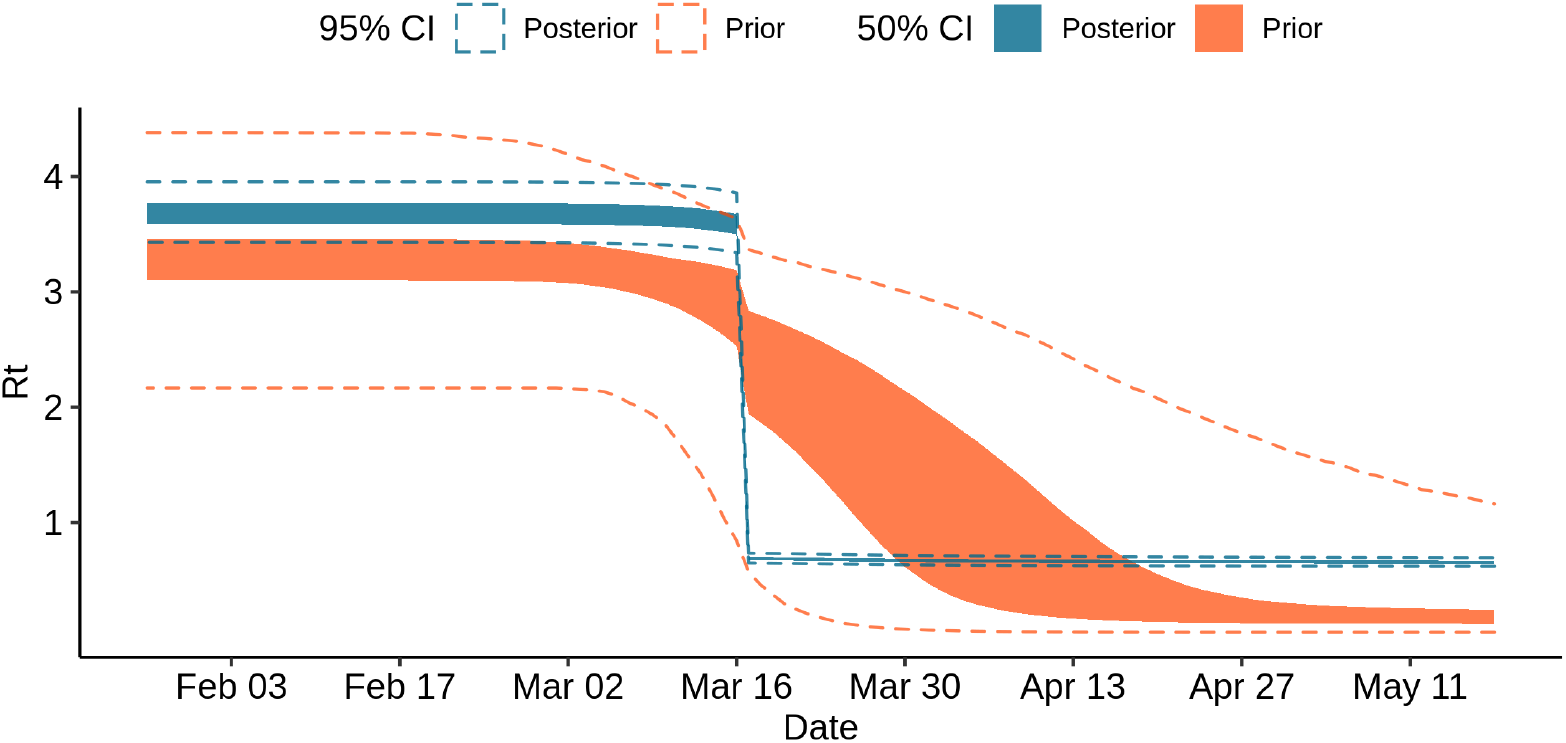
Prior and posterior samples of *R_t_* in region Ile-de-France.

### Effect of week-ends

Model 2 combines the effects of the lockdown and of week-ends. First we investigated what effect size would be necessary to detect an effect of week-ends on viral transmissibility, and then we assessed whether week-ends had had a detectable impact on viral transmissibility.

#### Effect size required to observe an effect of week-ends

Fig. 6 shows the effect on mortality in Ile-de-France through time and total mortality at national scale of decreases in *R_t_* due to a reduction of contacts between individuals on weekends, when fewer workers are active. They reveal that a *R_t_* fold change of around 0.75 seems necessary for it to have a detectable impact on the number of deaths, because the distributions obtained with an *R_t_* fold change of 0.9 overlap largely with the distributions obtained without a fold change. In terms of contacts, this would mean that there should be 25% fewer contacts during week-ends than during a week-day for the effect to be detectable. Simulation results for all regions are available as supplementary material.

**Figure 6.**
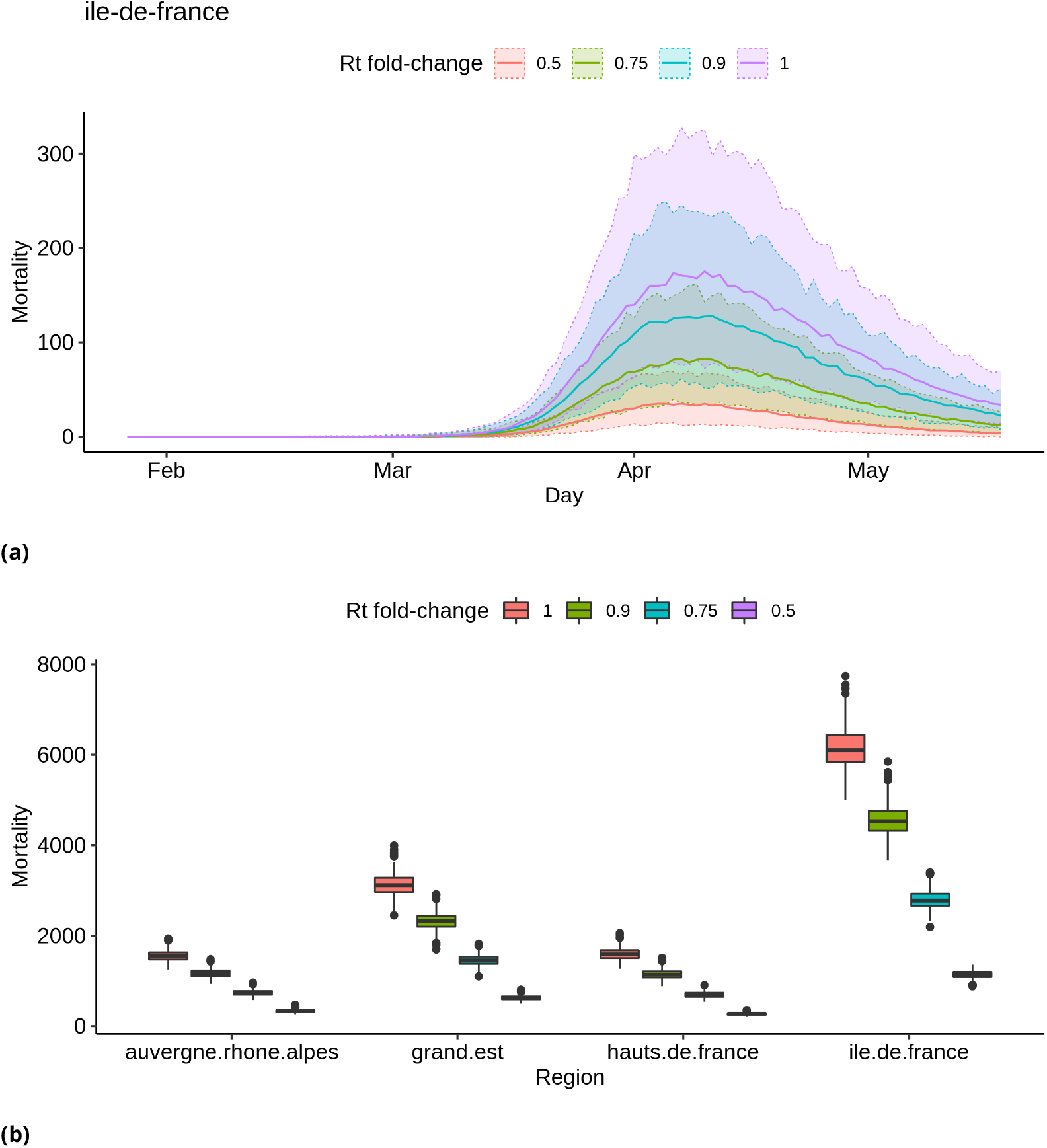
Simulated distributions of deaths, assuming different effect sizes of week-ends on *R_t_*. (a) Simulated distribution of deaths through time in region Ile-de-France. Median values are represented with a solid line, and shaded areas correspond to 95% credibility intervals. (b) Distributions of the total numbers of deaths in four regions. Each box shows (from top to bottom) the 3rd quartile, median and 1st quartile of the distribution. The vertical line on top of each box extends up to the largest value of the sample no further from the 3rd quartile than 1.5 times the inter-quartile difference; larger values are then represented as dots and can be interpreted as possible outliers. The vertical lines below each box are constructed in an analogous way for low values.

#### No detectable effect of week-ends on viral spread

The model finds little effect of changes of individual behaviour on week-ends on the dynamics of the number of deaths through time. Fig. 7 shows that the resulting posterior of *R_t_* looks very similar to the posterior obtained without accounting for behavioural changes on weekends (see Supplementary Figure 14 for comparison with base model *R_t_*). The associated posterior distribution of *α_weekend_* is presented in Supplementary Figure 12.

**Figure 7.**
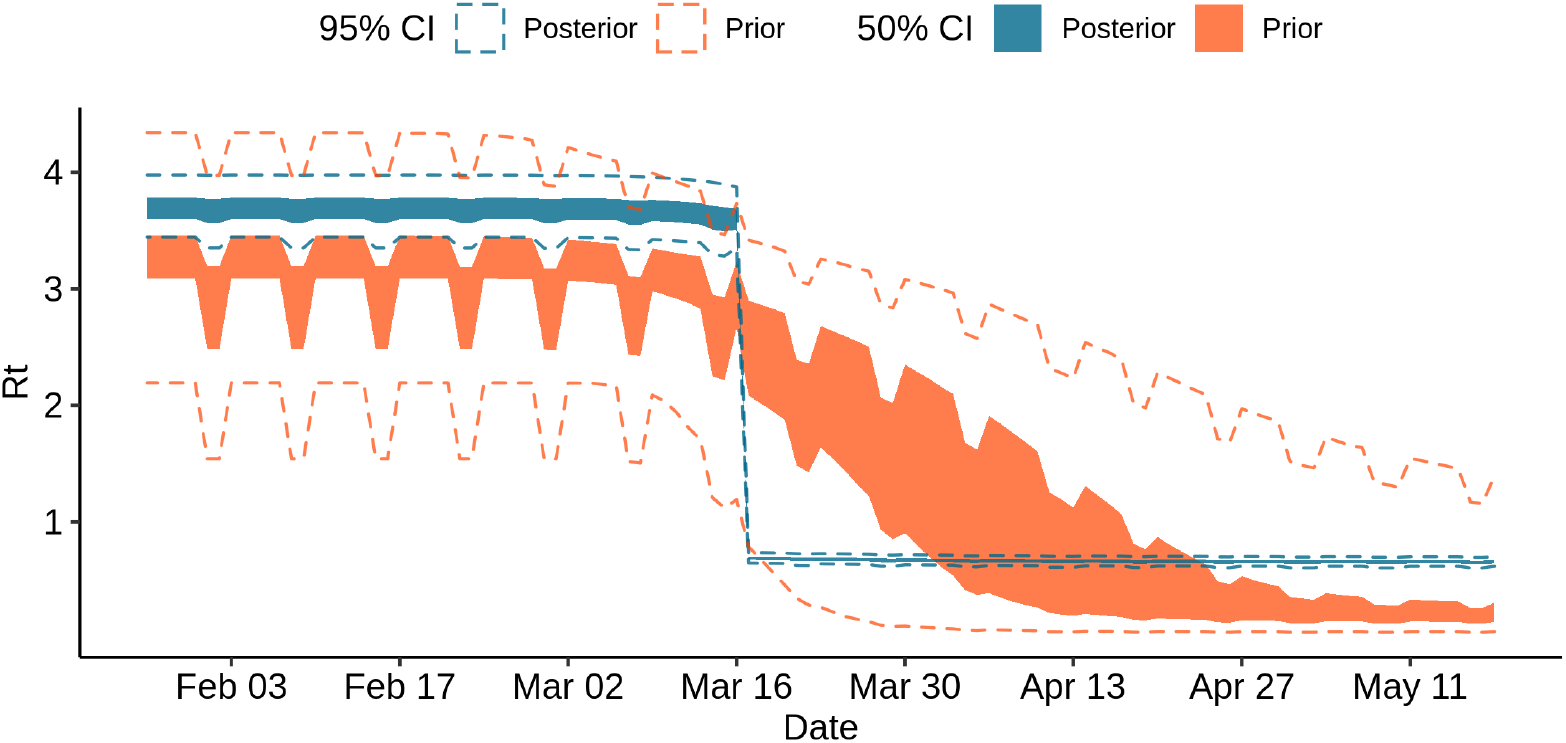
Prior and posterior samples of *R_t_* in region Ile-de-France

### Effect of the elections

The first round of voting in the municipal elections took place on Sunday March 15, just two days before the nation-wide lockdown was enacted. The voter turnout amounted to 41.6%. Following measures were enforced: safety distancing, and a maximum of three voters were allowed at once in polling stations; hydroalcoholic gel was available in every polling station, and masks were mandatory; voters were encouraged to bring their own pen and ballot paper which was sometimes sent by mail. Even with these precautions, such an event is expected to increase the number of contacts that occurs during the day, as well as the reproduction rate.

Model 3 combines the effects of the lockdown and of the election day. First we investigated what effect size would be necessary to detect an effect of the election day on viral transmissibility. Using simulations, we investigated different fold change values for the *R_t_* parameter. Second, we assessed whether the election day had had a detectable impact on viral transmissibility using the French mortality data.

#### Effect size required to observe an effect of the election day

Fig. 8 suggest that in order to detect an increase of the transmission rate *R_t_* on the election day based on mortality data, this effect would have to be a change in *R_t_* of at least a factor 2. This suggests that a model based of the number of deaths through time could only detect strong increases of *R_t_* during the election day. Additional simulation results for all regions are presented as supplementary material.

**Figure 8.**
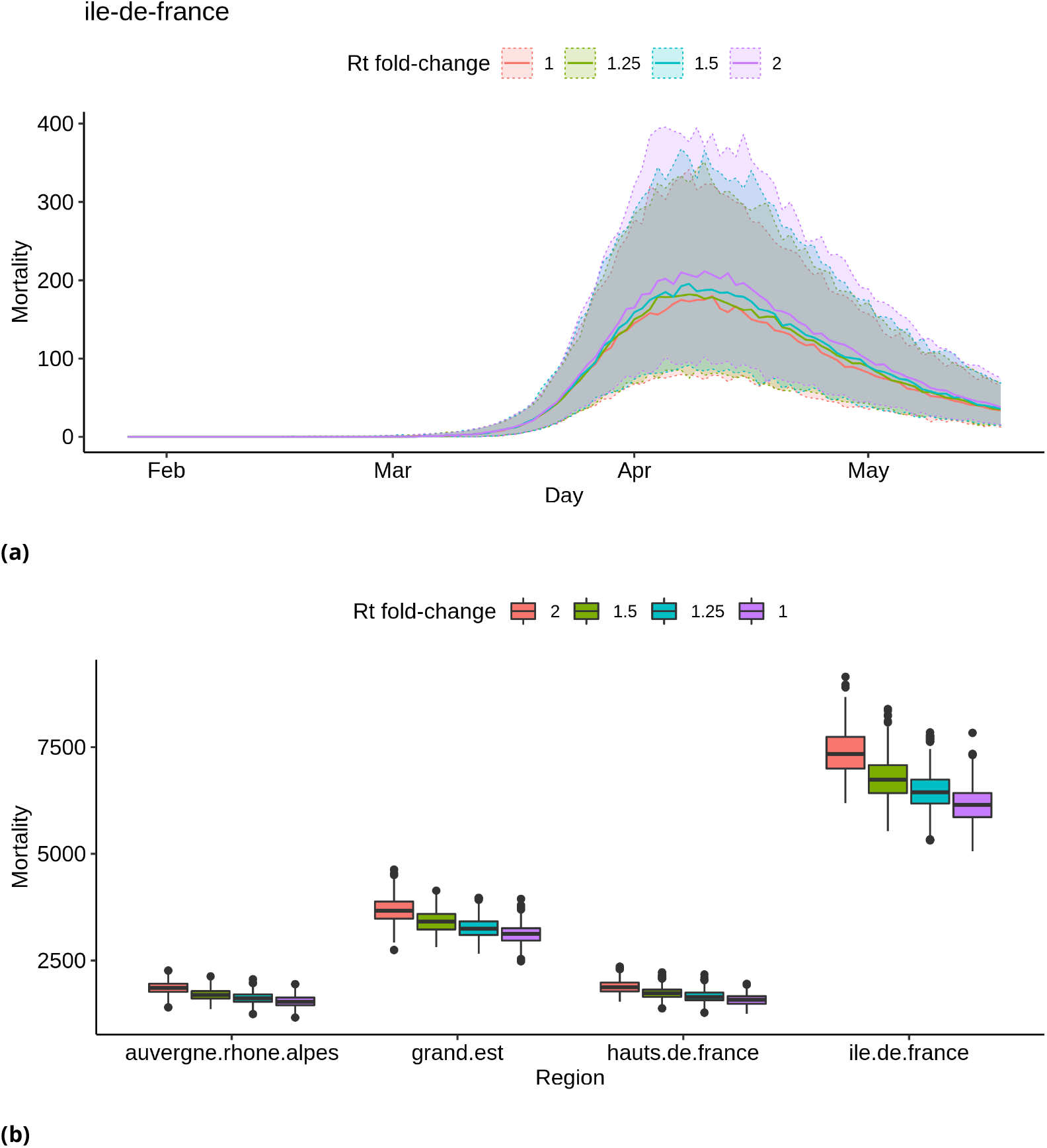
Simulated distributions of deaths count, assuming different effect sizes of the election day on *R_t_*. (a) Simulated distribution of deaths through time in region Ile-de-France. Median values are represented with a solid line, and shaded areas correspond to 95% credibility intervals. (b) Distributions of the total numbers of deaths in four regions. See Figure 6b for details on the representation.

#### No detectable effect of the election day on viral spread

The model finds no evidence for an increase in the number of contacts during election day on the dynamics of the number of deaths through time. Fig. 9 shows that the resulting posterior on the *R_t_* value is much flatter on March 15 than the prior. The associated posterior distribution of *α_elections_* is presented in Supplementary Figure 13.

**Figure 9.**
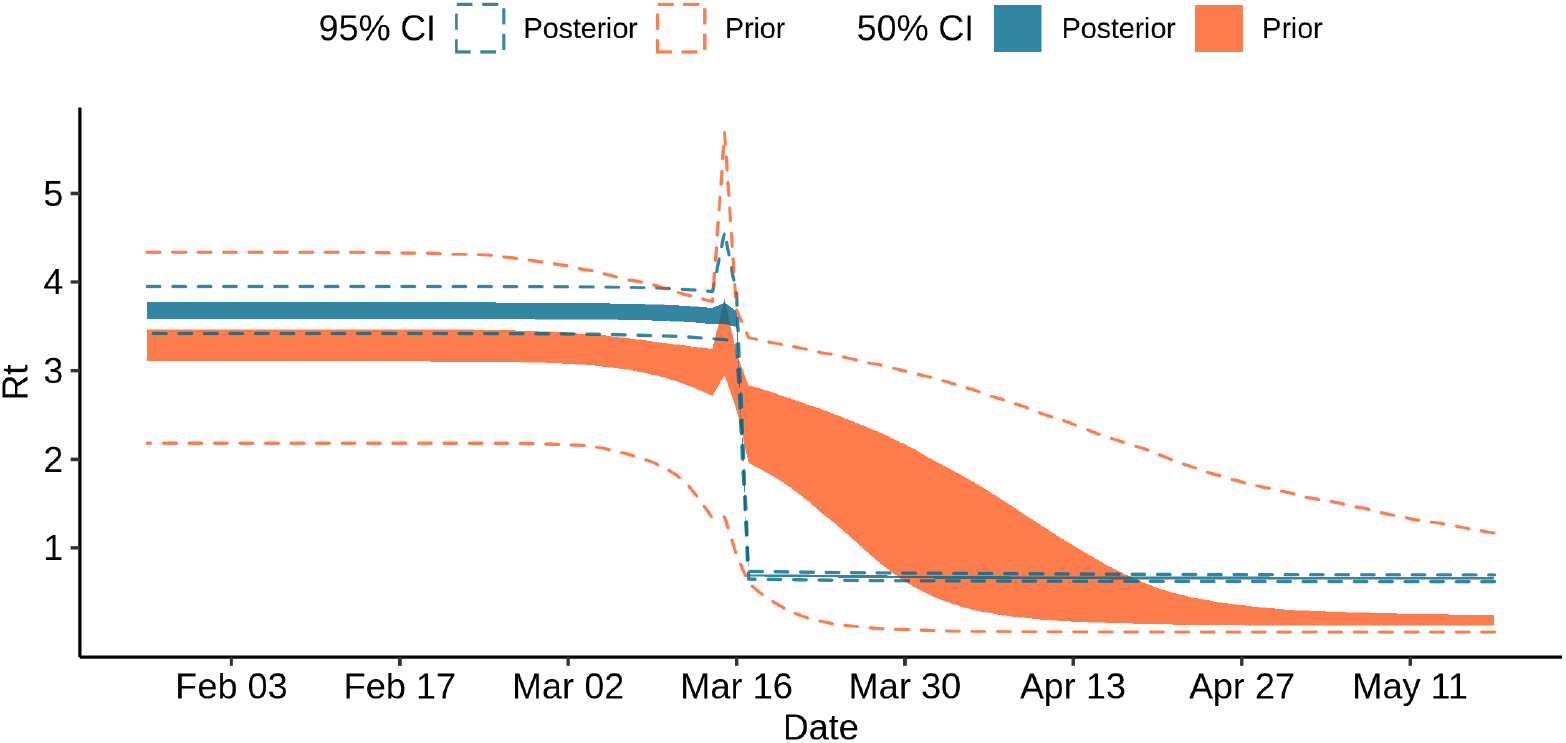
Prior and posterior samples of *R_t_* in region Ile-de-France

### Evidence for heterogeneity between regions in the efficacy of the lockdown

It has been suggested that the lockdown may have not been applied as strictly in different French regions. To investigate this, we used a mixture model to allow for two categories of reduction of the transmissibility due to the lockdown. We estimated two *α_lockdown_* values, one for each category of the mixture, and estimated a proportion *θ_i_* associated to each category. We found that the two categories almost had the same share among the 13 regions, with *θ*_1_ = 0.52 and *θ*_2_ = 0.48; comparison between the prior and the posterior distributions indicates that the data informed the model (Supplementary Figure 7). The corresponding reduction factors were 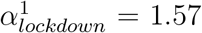 and (95 %C.I. 1.46 - 1.65) and 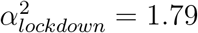 (95 %C.I. 1.67 - 1.94). We used posterior decoding to assign to each region a distribution of the *R_t_* fold change due to the lockdown (Fig. 10), defined as *exp*(−*α*) in equation 1. The distributions appear to be bimodal, which is expected given the underlying two categories of *α_lockdown_* used in the mixture model. The sizes of the modes vary depending on the region, which reveals that the two *α_lockdown_* values fit the regions differently. The lower *R_t_* fold change fits best the regions Île de France or Corse, while the higher *R_t_* fold change fits best Hauts de France and Occitanie.

**Figure 10.**
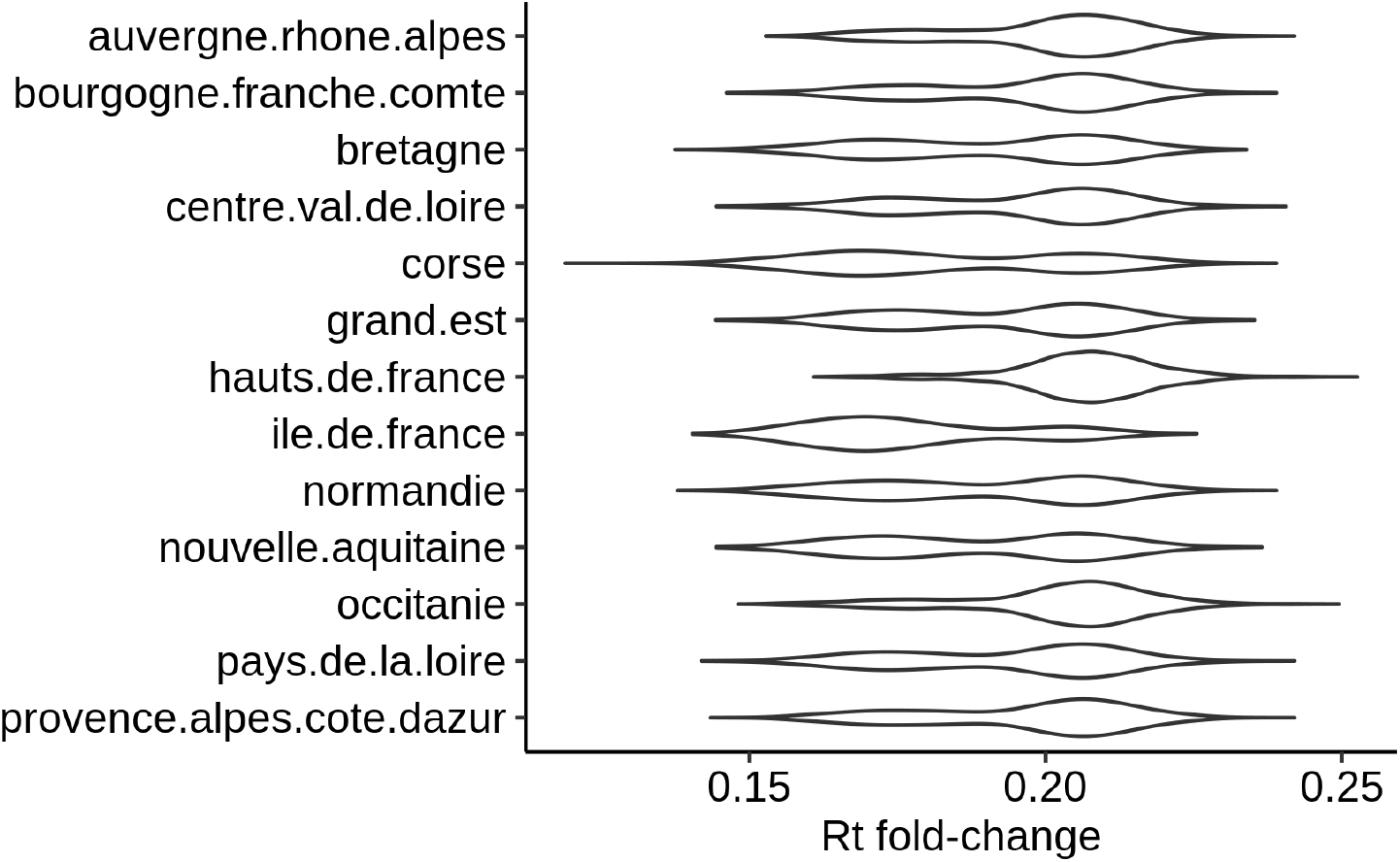
Posterior distribution of *R_t_* fold change per region

Median fold changes vary between 0.174 for Île de France and 0.207 for Hauts de France. Île de France is the region where the lockdown has had the strongest effect on the *R_t_*, contrary to expectations based on news reports. We used a linear model to investigate the relations between *R_t_* fold change as a dependent variable and regional population size, population density, and difference between pre- and post-lockdown population sizes as explanatory variables. This difference between pre- and post-lockdown population sizes is due to migrations between regions during the few days surrounding the lockdown decree. Our linear model has an adjusted *R*^2^ of 0.45. For each variable included in the model, we asked whether the corresponding coefficient in the linear regression was significantly different from 0. The most significant association we found was with population density, with a p-value of 0.02 and a negative correlation.

We compared the adjustement of the mixture model compared to that of model 1 by computing sums of squared errors over each day up to May 11. Squared errors are calculated for each sample between daily numbers of deaths and the numbers of deaths as predicted by each model. We found that the mixture model has a smaller error at 257950 (95% CI: 193776-345351) than model 1 at 283397 (95% CI: 211504-379692), representing a reduction of about 9% (Supplementary Figure 9). The reduction in error made by using a mixture model also varies depending on the region (Supplementary Figure 10), with the largest improvement observed in Île de France. There is support in the data for using a mixture model as shown by the difference between posterior and prior distributions (Supplementary Figure 7).

However, predictions on the last week of data when fitting on the corresponding prefix of data are not enhanced through the mixture model with total squared error equal 12975 (95% CI: 7335; 34699) when compared to model 1 (12350 [95% CI: 7051; 25307]). A more thorough evaluation of prediction performances, such as cross-validation, would be necessary to conclude on the general predictive capacity of both models.

The estimates of the national average reproduction number according to the mixture model are 3.25 (95% CI: 3.10 - 3.44) before lockdown and 0.63 (95% CI: 0.59 - 0.67) after.

### Status of the epidemic on May 11

We used both the mixture model and model 1 to assess the status of the epidemic on May 11 the day before the lockdown was lifted. Model 1 estimates that on May 11 2.09 (95% CI: 1.69-2.66) million people had been infected. This represents 3.22% (95% CI: 2.61-4.09) of the population. Further, the model estimates that there were 2793 (95% CI: 1761-4543) new infections on May 11.

The mixture model estimates that until this date 2.08 (95% CI: 1.85-2.47) million people had been infected, representing 3.20% (95% CI: 2.85-3.81) of the population. According to this model there were 2567 (95% CI: 1781-5182) new infections on May 11.

### Counterfactual investigation of alternative lockdown enforcements

We used our models to investigate the effect of putting the lockdown in place either earlier or later than the actual lockdown date on March 17. To do so, we assessed the total number of deaths predicted by the model as of May 11, a quantity that is well estimated by model 1 and by the mixture model as seen on Fig. 4. For the mixture model, Fig. 11 shows that delays in starting the lockdown result in excess deaths: from 21% (3575) additional deaths for one day of delay to 266% (45932) for 7 days of delay. Conversely, an earlier lockdown results in lower numbers of deaths, 76% (13044) fewer deaths for 7 days, and 19% (3204) for one day. For model 1, the trend is very similar with respectively: 21% (3666), 262% (45172), 75% (12997), 18% (3098).

**Figure 11.**
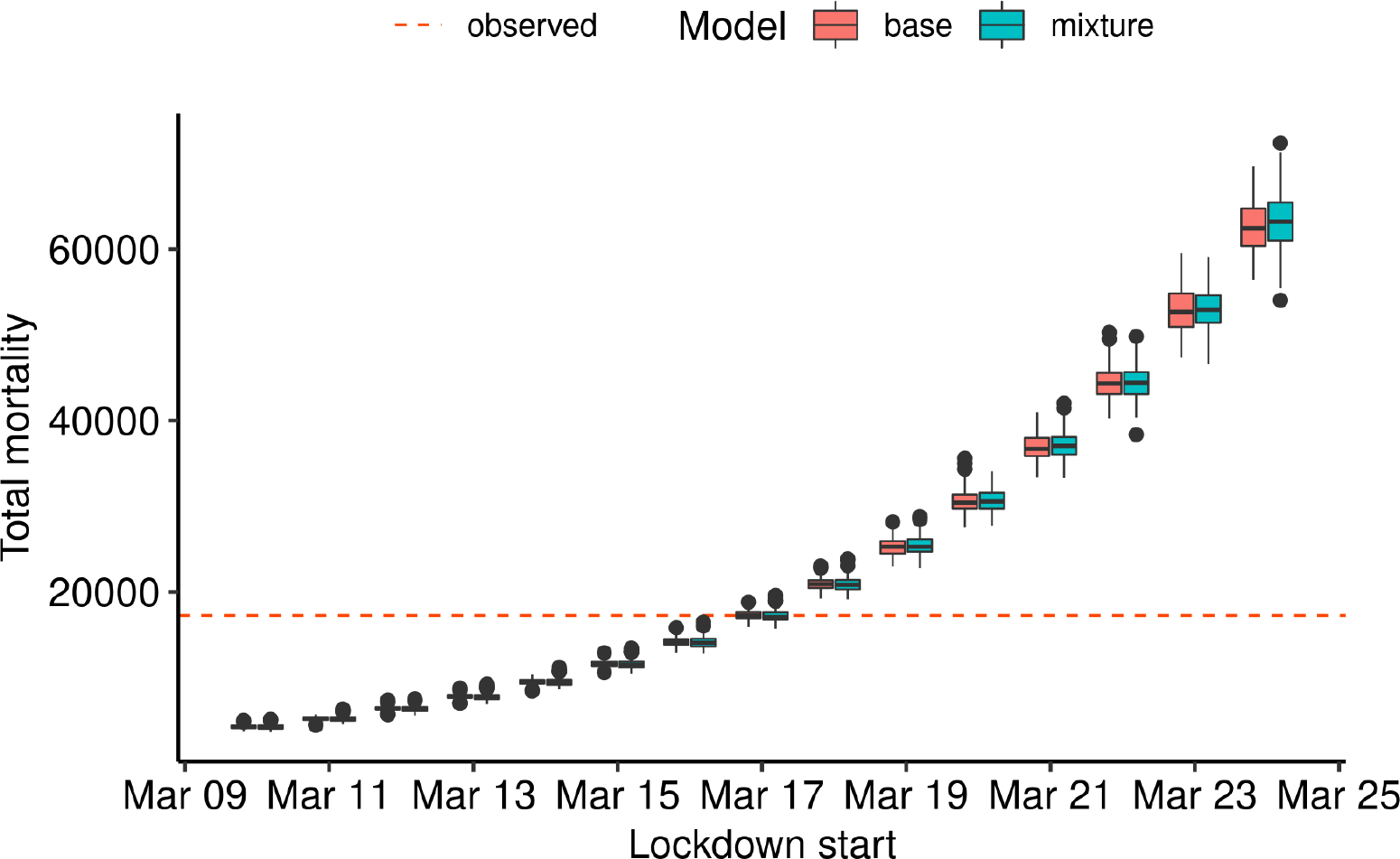
Effect of different lockdown dates in counterfactual scenarios. Both models were used to predict the total number of deaths on May 11 if the lockdown was put in place up to 7 days before or 7 days after the actual lockdown date on March 17.

## Discussion

In this manuscript, we studied the ability of a Bayesian model to fit the mortality data of the SARS-CoV-2 epidemic in France. These mortality data are incomplete, as they only include the numbers of deaths in hospitals of patients positive for the virus. In particular, they do not include deaths at home, or deaths in retirement facilities. Such input data also neglect other potentially useful sources of information, such as the number of cases, or the number of hospitalizations. Despite their shortcomings, numbers of deaths in hospitals have been widely used to study the epidemic in France and in other countries as it unfolded, notably because they were more readily available than other statistics.

We assessed the ability of our model to predict the number of deaths based on censoring of the data, and found that the model was able to accurately predict the number of deaths weeks in advance (Fig. 2).

We further explored the ability of our model using solely the number of deaths through time to detect the effect of week-ends or of single-day events, such as the election day. Weekends would need to incur a decrease of about 20% in e.g. the number of contacts to be detectable by the model. This was not found in the empirical data. The difference between weekdays and week-end days is probably weaker during lockdown, because fewer people go to work on any day during the lockdown. A single-day event would need to e.g. multiply the number of contacts on that day by a factor of 2 to be detectable; expectedly, the model found no evidence for such a large effect of the elections on the number of deaths. Accordingly, another study using admissions and deaths together with regional participation to the election has also found an absence of evidence that the elections had had a detectable impact on viral spread (Zeitoun et al., 2020).

We investigated whether the lockdown had had different effects on the reproduction number in the 13 French regions. Our mixture model identified differences between regions, with Ile de France showing the largest effect of the lockdown. This heterogeneity is not significantly correlated to differences in pre-lockdown R_0_, population sizes, areas, or the difference between the number of inhabitants pre and post lockdown. However, it is weakly negatively correlated to population density: the lockdown tends to be more efficient in denser regions.

Estimates obtained with the mixture model differ slightly from those obtained with model 1. For instance, nationally the average reproduction number is a bit smaller before and after lockdown (3.25 vs 3.34, and 0.62 vs 0.65). These estimates of the reproduction number can be compared to the values estimated by other groups. We focus on two works: those of (Salje et al., 2020) and (Sofonea et al., 2020).

(Salje et al., 2020) and (Sofonea et al., 2020) found results that are a bit different from ours, in particular for the reproduction number before the lockdown. The former estimated a reproduction number of 2.90 (95% CI:2.80-2.99) before the lockdown, and of 0.67 (95% CI:0.65-0.68) after the lockdown, and the latter a reproduction number of 2.99 (95% likelihood interval 2.59-3.39), and “between 21.3 and 27.1% of its value after the lockdown”, *i.e*. between 0.64 and 0.81. Our credibility intervals thus overlap with the intervals of (Sofonea et al., 2020). This is interesting as (Sofonea et al., 2020) used a different model from ours, that did not take into account heterogeneities between regions, but that is based on a probabilistic fine-grain compartmental model. (Salje et al., 2020) used a Bayesian model similar to ours, except that they used both hospitalization and deaths data, but did not model the saturation of the population as the epidemic progresses and the proportion of susceptible individuals decreases in the population, and did not use a mixture model to account for heterogeneities in the lockdown efficacy between regions.

A source of difference between our model, the model of (Sofonea et al., 2020), and theirs is the values of the Infection Fatality Ratios that were used. They based their IFR on the data from the Diamond Princess cruise ship, while (Sofonea et al., 2020) and we based ours on data from Wuhan, in China. As a result, their average IFR, nation-wide, is 0.7, while ours is 0.99. We performed a test by scaling down our IFRs by multiplying them by 0.7/0.99 in model 1. We find reproduction numbers in our results are virtually unchanged by this scaling of the IFR.

Values of the reproduction number in turn affect the estimates of the total number of infected people and the total number of new infections on May 11. (Salje et al., 2020) estimate that 2.8 (range: 1.8-4.7) million people have been infected by May 11, when the lockdown was lifted, and that there were 3900 (range 2600-6300) new infections on May 11. A series of sensitivity analyses yielded a larger range of values, notably between 1700 and 9600 new infections on May 11. These values are consistent with our estimates of the number of new infections on May 11. However, the mixture model infers that only 2.08 million people had been infected by May 11 (vs 2.09 for model 1), with 2567 new infections (vs 2794 for model 1). The difference in the total number of infections with (Salje et al., 2020) is likely explained by our higher IFR: fewer infections are required to explain a given number of deaths. Indeed, down-scaling our IFRs resulted in an increase ofthe total numberof infections to 2.71 millions (95% CI: 2.19 - 3.49) as of May 10 for model 1, closer to the estimate reported by (Salje et al., 2020). Better estimates of regional IFRs might be obtained by updating the work of (Roques et al., 2020) with more data. However, the better fit of the mixture model over model 1 suggests that the total number of infections is probably overestimated by model 1 and by (Salje et al., 2020). Overall, this comparison with (Salje et al., 2020) and (Sofonea et al., 2020) suggests that the estimates of key parameters of the epidemic are similar across a range of models and data sources, even if they do not fully agree.

Our study of counterfactual scenarios suggests that imposing the lockdown early results in fewer deaths, and imposing the lockdown late results in more deaths, which is unsurprising given the dynamics of any epidemic. It can be put in perspective with our study ofthe effect ofthe elections on the French epidemic. Although holding the elections on Sunday March 15th did not leave a noticeable footprint in the number of deaths, it may have caused a delay in imposing the lockdown. For instance, and according to the projections of our mixture model, setting up the lockdown on Friday March 13 instead of Tuesday March 17 would have resulted in 50% fewer deaths nationwide (8557 fewer deaths as of May 11, while the estimate according to model 1 is 55% (9466 fewer deaths as of May 11)).

## Conclusion

We used Bayesian models ofthe number of SARS-CoV-2 related deaths through time to study the epidemic, assess the influence of various events, and evaluate counterfactual scenarios. We found that the model accurately predicts the number of deaths a few weeks in advance, and recovers estimates that are in agreement with recent models that rely on a different structure and different input data. We also found evidence for heterogeneity between regions in the efficacy ofthe lockdown on epidemic spread. The predictions ofthe model indicate that holding the elections on March 15 did not have a detectable impact on the total number of deaths, unless it motivated a delay in imposing the lockdown.

### Availability

The code used for the experiments is available at our Git repository: https://gitlab.in2p3.fr/boussau/corona_french_regions; an archive ofthe code state at the time of publication is available at https://zenodo.org/record/4019099. All data used in this manuscript can be downloaded using dedicated scripts in the repository.

## Data Availability

The code used for the experiments is available at https://gitlab.in2p3.fr/boussau/corona_french_regions and as an archive at https://zenodo.org/record/4019099

## Supplementary material

Supplementary material is available online: https://www.medrxiv.org/content/10.1101/2020.06.09.20126862v4.supplementary-material

## Acknowledgements

The authors would like to thank (Flaxman et al., 2020) for making their model implementation open-source, thus allowing us to extend and modify it for the purpose of this research. We would also like to thank Wayne Landis and an anonymous reviewer for their fruitful comments on an earlier version of the manuscript.

Version 5 of this preprint has been peer-reviewed and recommended by Peer Community In Mathematical & Computational Biology (https://doi.org/10.24072/pci.mcb.100001).

